# Relationship between fill volume and transport in peritoneal dialysis

**DOI:** 10.1101/2024.01.31.24302046

**Authors:** Carl M. Öberg

**Affiliations:** Department of Clinical Sciences Lund, Skåne University Hospital, Lund, SE-21185, SWEDEN

**Keywords:** Keywords, Peritoneal dialysis, Diffusion capacity

## Abstract

**Introduction:** Larger fill volumes in peritoneal dialysis (PD) typically improve small solute clearance and water removal, and *vice versa* – but the relationship between intra-peritoneal volume and the capacities for solute and water transport in PD has been little studied. Here, it is proposed that this relative relationship is described by a simple ratio (Volume_new_/Volume_old_)^2/3^ up to a critical break-point volume, beyond which further volume increase is less beneficial in terms of transport.

**Method:** To scrutinize this hypothesis, experiments were conducted in a rat model of PD alongside a retrospective analysis of clinical data from a prior study. Rats underwent PD with either three consecutive fills of 8+8+8 mL (n=10) or 12+12+12 mL (n=10), with 45-minute dwell time intervals. This approach yielded sixty estimations of water and solute transport, characterized by osmotic conductance to glucose (OCG) and solute diffusion capacities, respectively.

**Results:** Comparative analysis of the predictive efficacy of the two models — the simple ratio *versus* the break-point model — was performed using Monte Carlo cross-validation. The break-point model emerged as a superior predictor for both water and solute transport, demonstrating its capability to characterize both experimental and clinical data.

**Conclusion:** The present analysis indicates that relatively simple calculations can be used to approximate clinical effects on transport when prescribing a lower or higher fill volume to patients on PD.

## Introduction

The diffusion capacity is the maximal clearance across a membrane, and is a key parameter both for hemodialyzer membranes (abbreviated K_0_A) and the peritoneal membrane (commonly abbreviated MTAC or PS). The diffusion capacity is proportional to the membrane surface area, which means that the diffusion capacity in PD is dependent on the fill volume. One can express the surface area *A* of any physical object in terms of its volume *V*, as follows

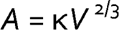

Here κ is a constant depending on the geometry (κ=1 for a cube and κ=6^2/3^π^1/3^ for a sphere *et cetera*). According to the square-cube law (Figure 1 A), changing the volume from *V*_1_ to *V*_2_ means changing the area from *A*_1_ to *A*_2_ as follows

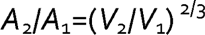

**Figure 1.**
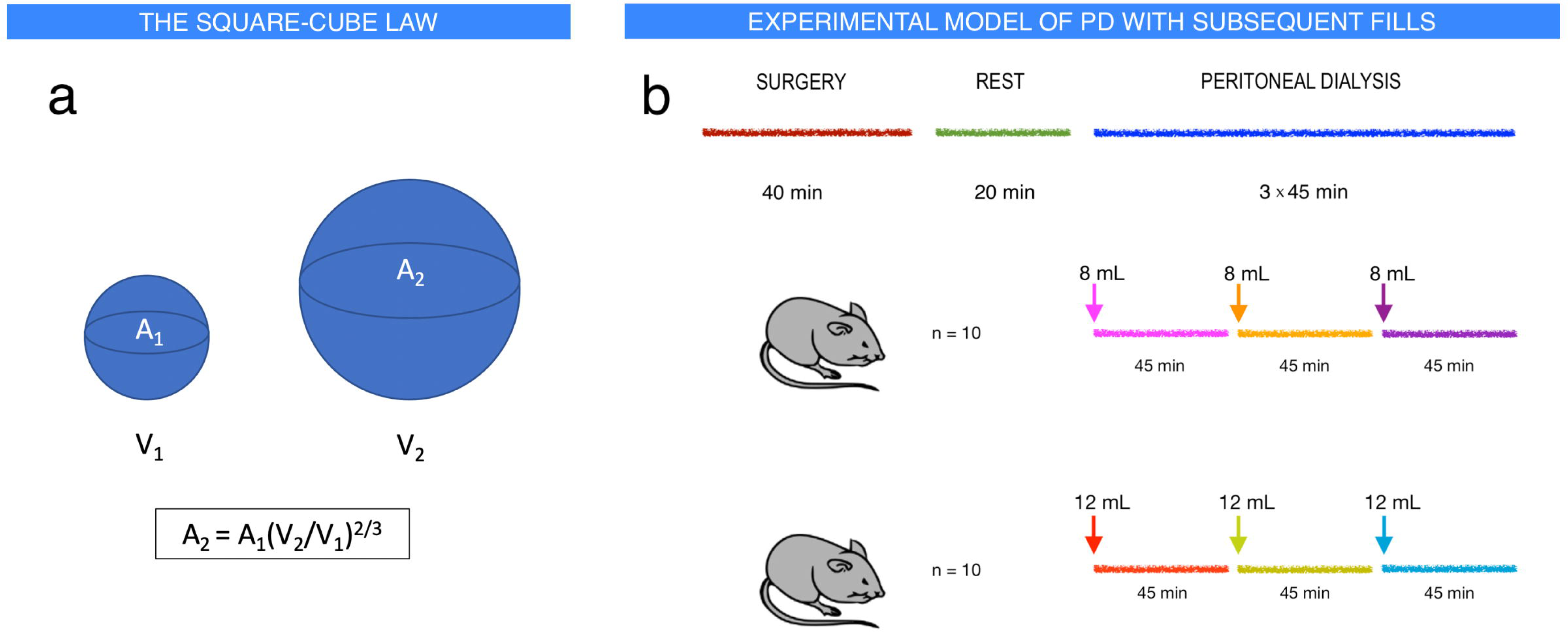
Schematic diagram of the physical square-cube law and the experimental setup. (**a**): The square-cube law in physics describes the relationship between the volume and the surface area as the volume of the object increases or decreases. The fractional increase or decrease in surface area is equal to the ratio of the volumes to the power of 2/3. Thus, decreasing the volume from 2,000 mL to 1,500 mL will reduce the surface area by 1 – (1500/2000)^2/3^ ≈ 17%. (**b**): Peritoneal dialysis was performed in twenty Sprague-Dawley rats. After surgery (see text), there was a resting period of 20 min before dialysis. Peritoneal dialysis was then performed with a fill volume of either 8 mL (*n*=10) or 12 mL (*n*=10) followed by a 45 min dwell after which an additional fill (8 mL or 12 mL, same as initial fill) was performed followed by another 45 min dwell after which the last fill of 8 mL or 12 mL was performed followed by a final dwell period of 45 min. During each dwell, sampling of the dialysate was performed directly after fill, and at 15-, 30-, and 45-min. Blood samples were collected before and after dialysis.

If the peritoneal surface area has approximately equal capacity for diffusion per unit area (mL/min per cm^2^), one may simply convert a known diffusion capacity (MTAC) from one volume to another

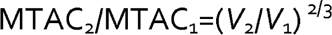

Relationships between fill volume and transport in PD have only rarely been studied (1, 2). In a study by Keshaviah and colleagues (2) it was found that small solute diffusion capacities appeared to reach a plateau phase or even decrease at higher fill volumes – apparently with a break-point volume depending on the body surface area. The authors however used phenomenological models instead of the physical square-cube law to describe the relation between fill volumes and solute transport. Moreover, the relationship between water permeability (*e.g.,* osmotic conductance) and fill volume was not assessed.

In this article, two competing models (square-cube law only *versus.* break-point model) for the relationship between fill volume and transport are evaluated, first in an experimental rat model of PD and, secondly, by analyzing clinical data from (2).

## METHODS

### Animals

Peritoneal dialysis was performed in twenty male Sprague-Dawley rats having an average body weight of 358 g (IQR 347 to 374). Prior to experiments all animals had free access to water and standard chow (Special Diets Services RM1(P) IRR.25 no. 801157). All animals were treated in accordance with the guidelines of the National Institutes of Health for Care and Use of Laboratory animals. The experiments were approved by the Ethics Committee for Animal Research at Lund University (Dnr 5.8.18-08386/2022). Results are reported in line with the ARRIVE guidelines. Exclusion criteria were bleeding into the dialysate, death, circulatory failure, or respiratory failure as judged by the technician performing the experiments (H. Axelberg).

### Anesthesia and surgery

Induction was achieved by gently placing the rat into a covered glass cylinder to which a continuous flow of 5% isoflurane in air (Isoban, Abbot Stockholm, Sweden) was connected. When fully anesthetized, the rat was removed from the container while anesthesia was maintained using 1.6-1.8% isoflurane in air delivered in a small mask. A tracheostomy was then performed via which the rat was connected to a volume-controlled ventilator (Ugo Basile; Biological Research Apparatus, Comerio, Italy) utilizing a positive end-expiratory pressure of 4 cm H_2_O. End-tidal *p*CO_2_ was kept between 4.8 and 5.5 kPa (Capstar-100, CWE, Ardmore, Pa). Body temperature was kept between 37.1°C to 37.3°C by using a feedback-controlled heating pad. The right femoral artery was cannulated and used for continuous monitoring of mean arterial pressure (MAP) and heart rate; and for obtaining blood samples (95 μL) for measurement of glucose, urea, electrolytes, hemoglobin, and hematocrit (I-STAT, Abbott, Abbott Park, IL), and blood ^51^Cr-EDTA activity. The right femoral vein was cannulated and connected to the dialyzer using plastic tubing. The right femoral artery was also cannulated and connected both to a pressure transducer to continuously monitor arterial line pressure. The right internal jugular vein was cannulated for infusion of maintenance fluid 3 ml/h. Hematocrit was determined by centrifugating thin capillary glass tubes. After the experiment, animals were euthanized with an intravenous bolus injection of potassium chloride.

### Experimental protocol

The experimental study consisted of twenty rats treated with 1.5 % glucose peritoneal dialysis fluid (Balance, Fresenius, Bad Homburg, Germany). Peritoneal dialysis was performed with three subsequent fills of either 8+8+8 mL (n=10) or 12+12+12 mL (n=10) separated by 45 min dwell time periods (Figure 1 B). For each dwell period, samples for analysis of ^51^Cr-EDTA and ^125^I-albumin activities were obtained from dialysate directly after instillation, at 15 min, 30 min, and after 45 min dwell time; and from plasma at 10 min, 20 min and 40 min dwell time. After each filling, dialysate samples for glucose concentration and electrolyte content (CHEM8) were obtained directly after fill and after 45 minutes. Arterial blood samples for routine chemistry (CHEM8) were collected before and after dialysis. Dialysate osmolality was assessed using the freeze-point method (Micro-Osmometer Model 210; Fiske Associates, Norwood, Massachusetts).

### Extraction and analysis of clinical data from Keshaviah et al

Average diffusion capacities and standard deviations (SDs) were obtained directly (Table 1 in (2)) while UF volumes and SDs were captured using WebPlotDigitizer (Figure 3 in (2)). OCG was estimated using the method by Martus *et al* (3) on the basis of measured ultrafiltration and D/D_0_ glucose. Since individual data points were not available, I resampled data using bootstrapping to generate 100 datasets on the basis of mean values and standard deviations in the original data. Statistical parameters were estimated from the arithmetic mean of the bootstrapped samples. Monte Carlo cross validation (4) was performed by randomly splitting each dataset (in a 60%:40% ratio) into 100 training sets and 100 validation sets. In the next step, competing models were fitted using segmented linear regression (see Supplemental material) to the training sets and evaluated (benchmarked) on the validation sets in terms of the root mean squared error – resulting in a total of 100 × 100 = 10,000 model comparisons.

**Table 1.**
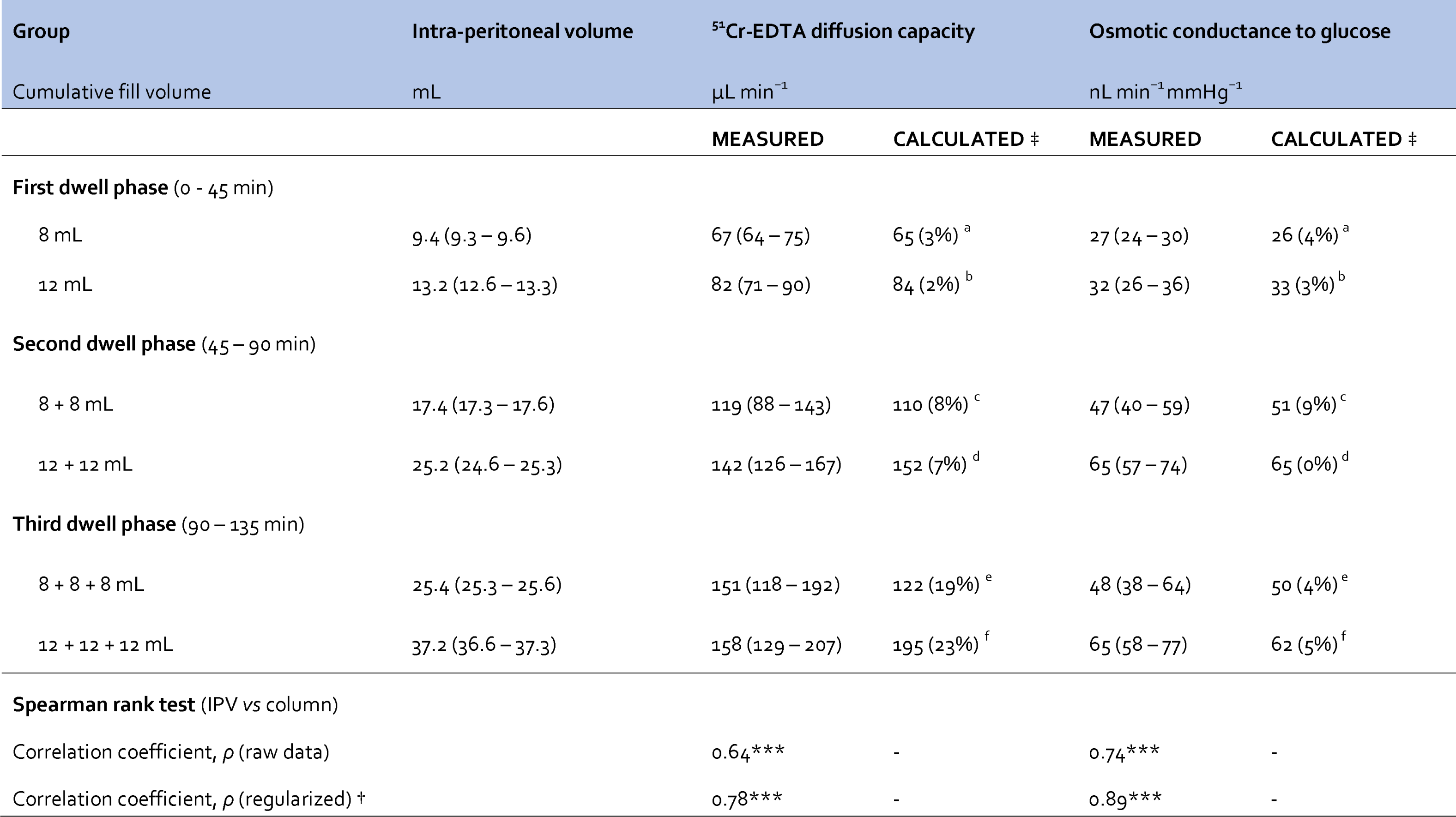
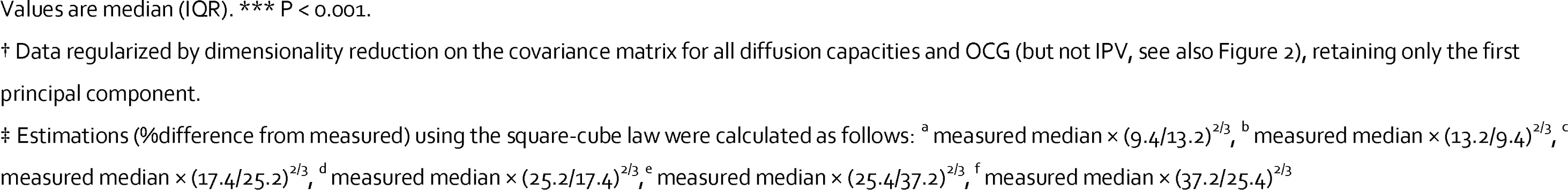
Intra-peritoneal volume *versus* solute and water transport capacities in Sprague-Dawley rats.

| Group | Intra-peritoneal volume | <sup>51</sup>Cr-EDTA diffusion capacity |  | Osmotic conductance to glucose |  |
| --- | --- | --- | --- | --- | --- |
| Cumulative fill volume | mL | μL min<sup>-1</sup> |  | nL min<sup>-1</sup> mmHg<sup>-1</sup> |  |
|  |  | MEASURED | CALCULATED ‡ | MEASURED | CALCULATED ‡ |
| First dwell phase (0 - 45 min) |  |  |  |  |  |
| 8 mL | 9.4 (9.3 – 9.6) | 67 (64 – 75) | 65 (3%)<sup>a</sup> | 27 (24 – 30) | 26 (4%)<sup>a</sup> |
| 12 mL | 13.2 (12.6 – 13.3) | 82 (71 – 90) | 84 (2%)<sup>b</sup> | 32 (26 – 36) | 33 (3%)<sup>b</sup> |
| Second dwell phase (45 – 90 min) |  |  |  |  |  |
| 8 + 8 mL | 17.4 (17.3 – 17.6) | 119 (88 – 143) | 110 (8%)<sup>c</sup> | 47 (40 – 59) | 51 (9%)<sup>c</sup> |
| 12 + 12 mL | 25.2 (24.6 – 25.3) | 142 (126 – 167) | 152 (7%)<sup>d</sup> | 65 (57 – 74) | 65 (0%)<sup>d</sup> |
| Third dwell phase (90 – 135 min) |  |  |  |  |  |
| 8 + 8 + 8 mL | 25.4 (25.3 – 25.6) | 151 (118 – 192) | 122 (19%)<sup>e</sup> | 48 (38 – 64) | 50 (4%)<sup>e</sup> |
| 12 + 12 + 12 mL | 37.2 (36.6 – 37.3) | 158 (129 – 207) | 195 (23%)<sup>f</sup> | 65 (58 – 77) | 62 (5%)<sup>f</sup> |
| Spearman rank test (IPV vs column) |  |  |  |  |  |
| Correlation coefficient, $\rho$ (raw data) | | 0.64*** | - | 0.74*** | - |
| Correlation coefficient, $\rho$ (regularized) † | | 0.78*** | - | 0.89*** | - |

Values are median (IQR). \*\*\* $P < 0.001$ .
† Data regularized by dimensionality reduction on the covariance matrix for all diffusion capacities and OCG (but not IPV, see also Figure 2), retaining only the first principal component.
‡ Estimations (%difference from measured) using the square-cube law were calculated as follows: <sup>a</sup> measured median $\times (9.4/13.2)^{2/3}$ , <sup>b</sup> measured median $\times (13.2/9.4)^{2/3}$ , <sup>c</sup> measured median $\times (17.4/25.2)^{2/3}$ , <sup>d</sup> measured median $\times (25.2/17.4)^{2/3}$ , <sup>e</sup> measured median $\times (25.4/37.2)^{2/3}$ , <sup>f</sup> measured median $\times (37.2/25.4)^{2/3}$

**Table 2.**
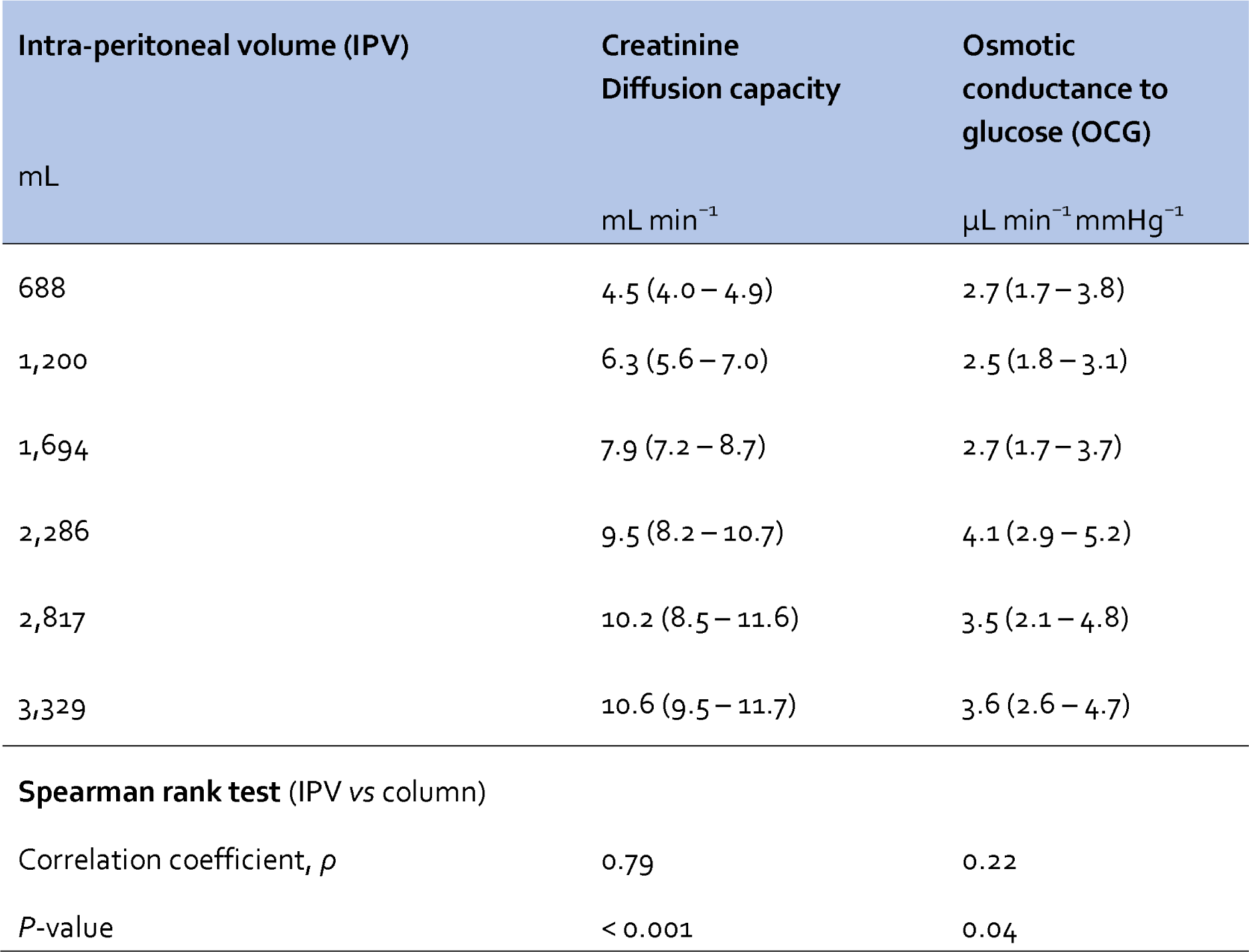
Intra-peritoneal volume *versus* solute and water transport capacities in patients.

### Calculations

Dialysate ^51^Cr-EDTA concentrations were fitted to an exponential model for each dwell period

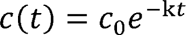

by doing linear regression on log-transformed data. Dialysate clearances were determined from where V is the intra-peritoneal volume estimated as the sum of filled volume and the residual volume estimated from the dilution of ^125^I-albumin, *i.e.* 8+rv or 12+rv for the first dwell period, 16+rv or 24+rv for the second dwell period, v ultrafiltration is thus neglected. Average ^51^Cr-EDTA concentration during each 45 min dwell period was determined from

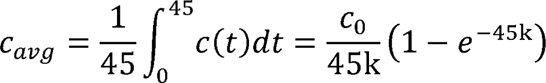

The residual volume was determined from the dilution of ^125^I human serum albumin (see e.g. (3)), ^51^Cr-EDTA and creatinine. For calculations I relied on the former to be most accurate. Ultrafiltration rates and osmotic conductance to glucose from each dwell period were estimated from sodium sieving using the method described in the recent article by Helman (5), using a counter-pressure of 25 mmHg which resulted in OCG values very similar to those obtained in Sprague-Dawley rats in a recent study (6). Small solute diffusion capacities were assessed using the isocratic method (see e.g. (7)).

### Statistical methods

Data are shown as median (interquartile range) unless otherwise specified. Correlations between transport parameters and intra-peritoneal/fill volumes were assessed using the Spearman’s rank test, which is a Pearson’s correlation test performed on ranked data. The p-value for the Spearman rank test was determined from the t-distribution. A simple power analysis (*pwr.r.test*, pwr package) showed that sixty measurements of diffusion capacity provided 80% power to detect an effect size (Spearman’s *ρ*) of at least 0.35. Significant differences between two groups were assessed using a non-parametric Wilcoxon test (wilcox.test) if not otherwise specified. P-values below 0.05 were considered significant. Non-linear regression was performed using the *nls* function in R. Calculations were performed using R for mac version 4.1.1.

## RESULTS

Experimental peritoneal dialysis was performed in twenty anesthetized Sprague-Dawley rats, with three subsequent fills of either 8+8+8 mL (n=10) or 12+12+12 mL (n=10) separated by 45 min dwell time periods, as shown in Figure 1 B. Routine- and treatment parameters are shown in Supplemental Table 1. No animal was excluded from analysis. As is seen in Table 1, calculated values using the square-cube law showed excellent agreement with measured values, especially for the first- and second dwell time periods.

### Dimensionality reduction using Principal Component Analysis

Principal component analysis (PCA) was performed on the correlation matrix of all measured transport parameters expected to correlate with peritoneal surface area (Figure 2 A). Dimensionality reduction (regularization) was achieved by retaining only the first principal component in the dataset, since most of the variance could be explained by the first principal component (Figure 2 B). Also, Horn’s parallel analysis showed that only the first component needed to be retained. The correlation between capacities for solute- and water transport and intra-peritoneal volume was markedly improved after regularization (Table 1 and Supplemental Table 2), supporting the hypothesis that the co-variance of the transport parameters in Figure 2 A is chiefly due to alterations in intra-peritoneal volume.

**Figure 2.**
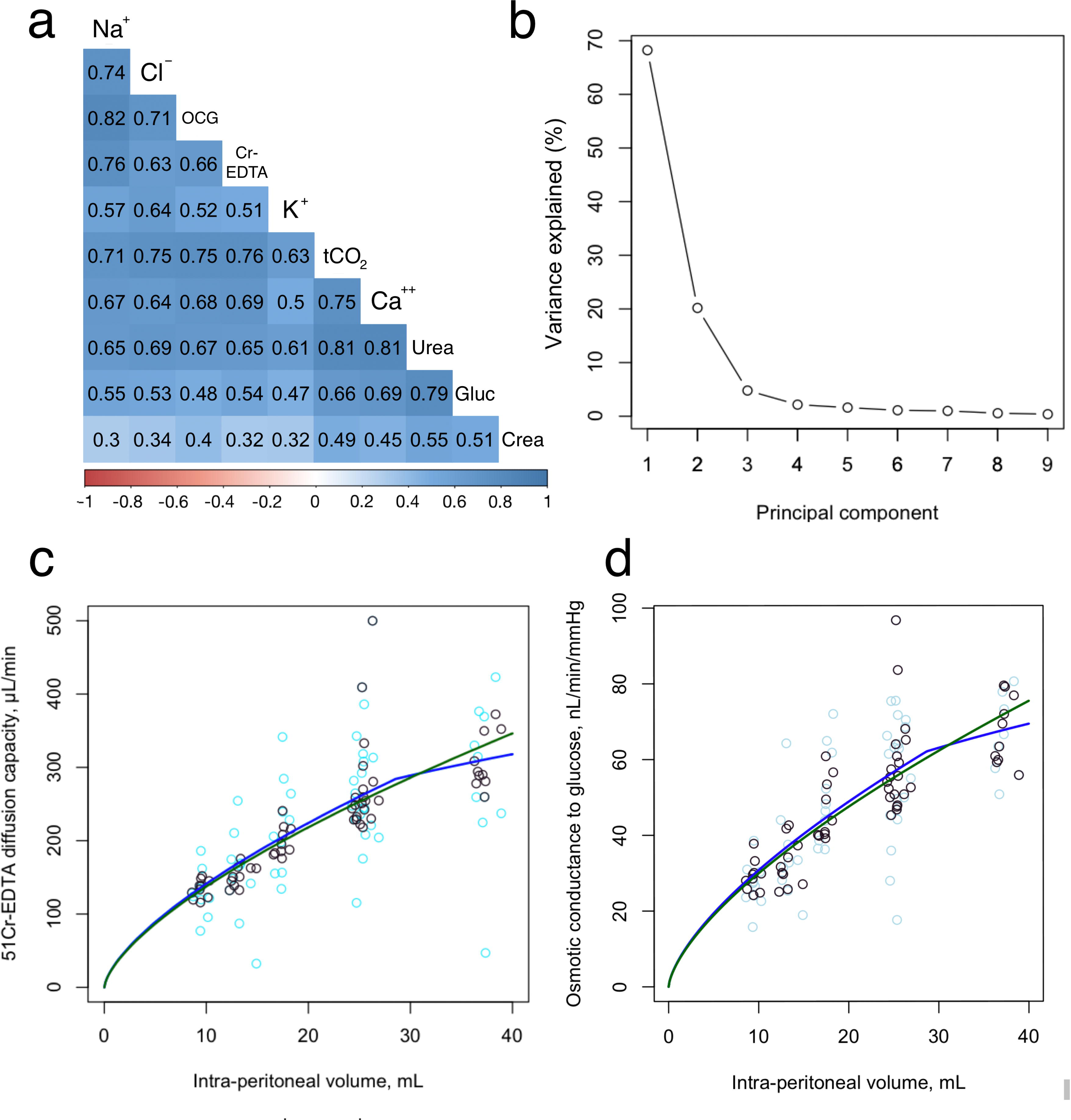
Correlation matrix between transport parameters in the rat model of PD, principal component) analysis and Monte-Carlo cross-validation of competing models. (**a**): Correlation plot between calculated diffusion capacities for sodium (Na+), chloride (Cl^−^), ^51^Cr-EDTA (Cr-EDTA), potassium (K+), total carbon dioxide (tCO_2_), calcium (Ca^++^), urea, glucose (Glu), creatinine (Crea), and osmotic conductance to glucose. Only significant correlations are displayed. (**b**): Scree-plot of the principal component analysis showing the %-explained variance per principal component. (**c**): Monte-Carlo cross validation comparing the ability of simple ratio model *versus* the break-point model to describe the relation between creatinine diffusion capacity (y-axis) and intra-peritoneal volume. (**d**): Monte-Carlo cross validation comparing the ability of simple ratio model *versus* the break-point model to describe the relation between osmotic conductance to glucose (y-axis) and intra-peritoneal volume. Analyses in (**c**) and (**d**) were performed both on raw data (light-blue circles) and regularized data (black circles) using dimensionality reduction (retaining only the first principal component).

**Figure 3.**
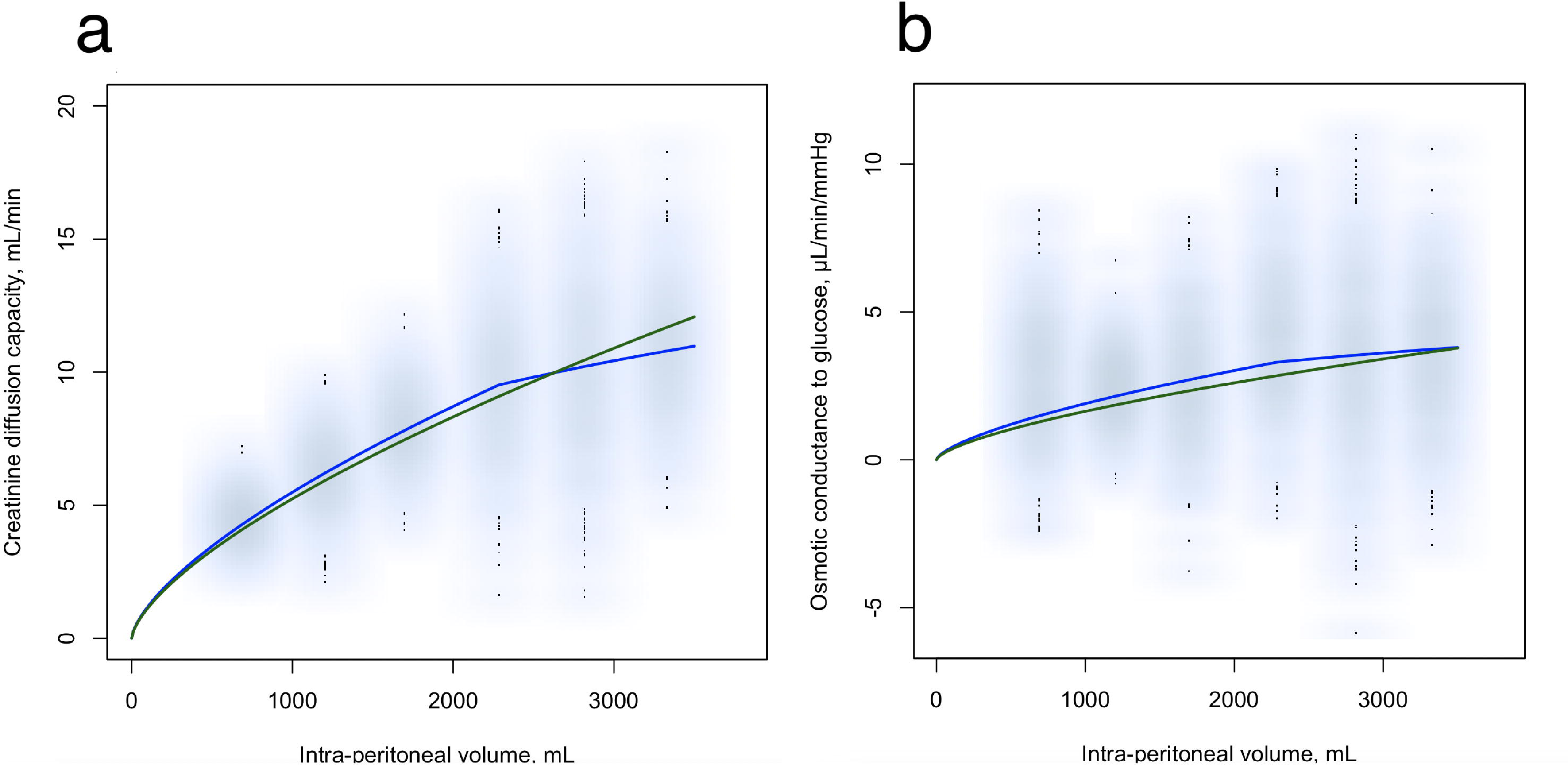
Comparative analysis of the predictive efficacy of the two models — the simple ratio versus the break-point model — performed using Monte Carlo cross-validation on clinical data. (**a**): Diffusion capacity for creatinine and (**b**): the osmotic conductance to glucose. Data was bootstrapped by re-sampling the dataset, resulting in 10,000 data-points plotted in light-blue color.

### Monte Carlo cross-validation shows that the breakpoint model provides a superior fit – both in experimental and clinical data

Next, the models were tested based on 100-fold Monte Carlo cross-validation with a 60%-40% train-test ratio. Regression was performed in each training set (36 datapoints) and the predictive capability of the model was thereafter assessed from the root mean squared error (RMSE) between the trained model and each validation dataset (24 datapoints). Results showed a better fit (lower RMSE) for the break-point model both for solute diffusion capacity and osmotic conductance to glucose (Supplemental Table 3). For the experimental rat data, the predicted MTACs for ^51^Cr-EDTA and OCGs according to the competing models are plotted in Figure 2 A and B along with regularized data (black rings) and non-regularized data (light-blue rings). Similarly, for the clinical data, predicted MTACs for creatinine and OCGs *versus* data are shown in Figure 3 A and B, respectively.

### Clinical implications on transport - applying Ohm’s law to Peritoneal Dialysis

The effects on OCG and MTAC by prescribing a different fill volume are easy to calculate using the above results. However, it is slightly more complex to assess the clinical effects on UF and parameters like the dialysate to plasma (or initial) solute concentration ratio. However, under the assumption of an exponential increase/decay of dialysate solute concentration, remarkably simple relationships may be derived using a variant of Ohm’s law (see Supplemental material). Thus, the time it takes for the dialysate concentration to reach a certain value in relation to the initial (or plasma) concentration (D/D_0_) is decreased/increased by the factor (V_new_/V_old_)^1/3^ when changing the fill volume to V_new_ from V_old_. This makes it possible to adapt the dwell time to preserve the osmotic gradient by using this factor, by shortening or prolonging dwell time by the factor (V_new_/V_old_)^1/3^. If the osmotic gradient is preserved in this manner, the trans-peritoneal UF rate is decreased/increased by the factor (V_new_/V_old_). For example, to achieve the same D/P-ratio for creatinine when lowering the intra-peritoneal volume from 2,000 mL to 1,000 mL, the dwell time needs to be shortened by 1-(1000/2000)^1/3^ ≈ 20%, and thus the transport status appears “faster” at a smaller fill volume. Such a reduction in dwell time will then lead to the trans-peritoneal UF volume being reduced by (1000/2000) = 50%. A few clinical scenarios were simulated using the three-pore model (Supplemental Table 4) modified by the use of the break-point model to scale MTACs and OCG. The simulations show excellent agreement with the factors for dwell time (V_new_/V_old_)^1/3^ and trans-peritoneal UF-volume (V_new_/V_old_).

## DISCUSSION

The present article provides simple and effective tools to predict effects on transport parameters when changing the fill volume from V_old_ to V_new_, using the simple approximation (V_new_/V_old_)^2/3^. It would be more accurate to use the intra-peritoneal volume rather than the fill volume. However, for typical fill- and residual volumes, taking this into account would only affect the factor by a few percent. Since the error of measurement is commonly larger than this in a clinical setting, taking RVs into account is thus unlikely to improve the estimate. For example, a decrease in fill volume from 2000 mL to 1000 mL will, on average, lead to an approximate decrease in small solute diffusion capacity and OCG by (1,000/2,000)^2/3^ ≈ 37%. If assuming a residual volume of 200 mL we instead get (1,200/2,200)^2/3^ ≈ 33%. Although the transport capacities for solutes and water are reduced by an equal amount, the relative decrease in diffusion capacity for the osmotic agent (glucose) is less than the relative decrease in volume (which is simply V_new_/V_old_). This, means that the peritoneum will appear “faster” when the fill volume is reduced since the clearance of the osmotic gradient will be higher in relation to the fill volume. *Vice versa* - by increasing the fill volume, the peritoneum will appear “slower”, and slightly longer dwell times may be used, according to the factor (V_new_/V_old_)^1/3^.

A strength of the present study is the fact that the same model was validated in both experimental data and clinical data, showing that the relationship between fill volume and the transport of water and solutes can be characterized using the square-cube law up to an average intra-peritoneal break-point volume (being ∼2,300 mL in patients, ∼26 mL in rats), after which a phenomenological line-cube equation provides a better estimate. The choice of a line-cube relationship is arbitrary, and most likely any monotonously increasing model would do. Instead, of using the square-cube law, phenomenological estimations have been used since long (8-10), which will provide very similar results. According to physical laws, alterations in transport (convection- and diffusion) capacities at different fill volumes should follow the square-cube law, so the present results are not surprising from a theoretical stand-point. Although the apparent transport capacity per unit surface area of the peritoneum may not remain constant when the intra-peritoneal volume changes, the present analysis convincingly shows that the square-cube law describes these changes well in the average sense. At higher intra-peritoneal volumes, it’s evident that the increments in convective- and diffusive transport capacities are less efficient compared to those at lower volumes. Several explanations are conceivable. Firstly, the increased intra-peritoneal pressure at higher volumes will likely act to reduce blood-flow across peritoneal tissues, reducing the apparent diffusion capacity. Higher intra-peritoneal pressures may also act to stretch the peritoneum so that the density of vessels per unit membrane area is lower at higher volumes. Regardless of the exact mechanism, the break-point volume is most likely unique for each patient. In fact, Keshaviah *et al (2)* showed a correlation between peak dialysate volume and body surface area, and it is conceivable that there is a similar correlation between break-point volume and body-surface area.

The present study has some limitations that affect the interpretation of the results. First, the calculation of diffusion capacity in rats was herein based on the assumption of a constant UF rate (isocratic model) during each 45 min dwell period. This is only approximately true for 1.5% glucose fluids, but previous work comparing isocratic diffusion capacities with the three-pore model (gold standard), showed excellent agreement (6). Another limitation is the use of bootstrapping to analyze clinical data, which relies on the assumption that the sampling distributions are normal. Indeed, it would be possible to just analyze mean values using non-linear regression, but here the bootstrapping approach is to be preferred. This is because such an approach would most likely provide a better fit for the break-point model due to the fact that it has one more parameter (break-point volume) compared to the reference square-cube model. Although corrections exist for such over-fitting, the approach using Monte Carlo cross-validation avoids this potential issue (4), since the models are validated on data that it has “never seen”.

The present results have clear implications on the prescription of high-quality PD, which nowadays is focused more on the individual patient (11), rather than on loosely defined treatment goals such as Kt/V urea. However, the main application of the present findings is to aid interpretation when performing peritoneal function tests using a different fill volume from the standard 2,000 mL (12). The corrections herein may also be applied when prescribing lower fill volumes, *e.g.* in incremental peritoneal dialysis (13) (14). Future clinical research could focus on the determination of the break-point volume in individual patients, while experimental studies could be performed to identify the mechanisms behind this “break-point” phenomenon.

## Data Availability

Original experimental data reported in this article have been deposited in figshare (doi: 10.6084/m9.figshare.24947982).

https://doi.org/10.6084/m9.figshare.24947982

## Acknowledgements

Helén Axelberg is gratefully acknowledged by the author for excellent experimental work. The study was funded by Lund University Medical Faculty Foundation grant YF 2020-YF0056, The Inga-Britt and Arne Lundberg’s Research Foundation, the Swedish Kidney Foundation, and the Crafoord Foundation (all to C.M. Öberg).

