## Supplemental Material for "Relationship between fill volume and transport in peritoneal dialysis"

Carl M. Öberg, MD, PhD [1]*

[1] Department of Clinical Sciences Lund, Skåne University Hospital, Lund, SE-21185, SWEDEN.

### SUPPLEMENTAL MATERIAL

**Supplemental Table 1 A.** Hemodynamic and routine lab parameters before and after dialysis.

| Parameter | Before dialysis | After dialysis |
|---|---|---|
| Mean arterial pressure, mmHg | 119 (110 – 128) | 103 (86 – 114) * |
| Heart rate | 387 (360 – 402) | 325 (310 – 345) *** |
| Plasma sodium, mmol/L | 135 (135 – 136) | 137 (137 – 138) *** |
| Plasma potassium, mmol/L | 4.5 (4.3 – 4.6) | 4.3 (3.9 – 4.4) ** |
| Plasma total $CO_2$, mmol/L | 26 (25 – 27) | 24 (23 – 24) *** |
| Plasma chloride, mmol/L | 99 (98 – 100) | 104 (103 – 105) *** |
| Plasma ionized calcium, mmol/L | 1.38 (1.34 – 1.40) | 1.40 (1.37 – 1.42) * |
| Blood hemoglobin, g/L | 143 (139 – 146) | 136 (133 – 139) *** |

Values are median (IQR). * $p < 0.05$, ** $p < 0.01$, ** $p < 0.01$, *** ** $p < 0.001$.

**Supplemental Table 1 B.** Treatment parameters.

| Parameter | Value |
|---|---|
| Dialysis fluid glucose, mmol/L [a] | 83.2 |
| Dialysis fluid sodium, mmol/L [a] | 134 |
| Dialysis fluid potassium, mmol/L [a] | 0 |
| Dialysis fluid chloride, mmol/L [a] | 100.5 |
| Dialysis fluid lactate, mmol/L [a] | 35 |
| Dialysis fluid calcium, mmol/L [a] | 1.25 |
| Dialysis fluid magnesium, mmol/L [a] | 0.5 |
| Total fluid filled (group 1), mL | 24 |
| Total fluid filled (group 2), mL | 36 |
| Total fluid drained (group 1), mL | 26.8 |
| Total fluid drained (group 2), mL | 39.6 |

[a] Nominal values from the manufacturer (Balance, Fresenius Medical Care, Bad Homburg, Germany).

**Supplemental Table 2**

| **Group**<br>Cumulative fill volume | **Intra-peritoneal volume (IPV)**<br>mL | **UF rate**<br>μL $min^{-1}$ | **Creatinine MTAC**<br>μL $min^{-1}$ | **Glucose MTAC**<br>μL $min^{-1}$ | **Potassium MTAC**<br>μL $min^{-1}$ | **tCO2 MTAC**<br>μL $min^{-1}$ |
|---|---|---|---|---|---|---|
| **First dwell phase** | | | | | | |
| 8 mL | 9.4 (9.3 – 9.6) | 15 (12 – 17) | 149 (139 – 174) | 108 (98 – 131) | 319 (293 – 409) | 191 (158 – 217) |
| 12 mL | 13.2 (12.6 – 13.3) | 19 (18 – 24) | 196 (155 – 221) | 122 (88 – 135) | 347 (333 – 412) | 213 (180 – 228) |
| **Second dwell phase** | | | | | | |
| 8+8 mL | 17.4 (17.3 – 17.6) | 21 (20 – 26) | 331 (238 – 476) | 146 (125 – 165) | 369 (342 – 459) | 302 (260 – 335) |
| 12+12 mL | 25.2 (24.6 – 25.3) | 32 (28 – 38) | 570 (320 – 725) | 194 (152 – 212) | 505 (468 – 595) | 375 (335 – 416) |
| **Third dwell phase** | | | | | | |
| 8+8+8 mL | 25.4 (25.3 – 25.6) | 17 (14 – 21) | 609 (503 – 886) | 210 (182 – 229) | 469 (442 – 523) | 336 (301 – 388) |
| 12+12+12 mL | 37.2 (36.6 – 37.3) | 27 (25 – 29) | 482 (436 – 891) | 242 (200 – 251) | 613 (510 – 702) | 512 (463 – 540) |

**Spearman rank test** (IPV *vs* column)

| | | | | | |
|---|---|---|---|---|---|
| Correlation coefficient, *ρ* | 0.45 | 0.77 | 0.69 | 0.69 | 0.89 |
| Correlation coefficient (regularized), *ρ* | 0.53 | 0.90 | 0.81 | 0.91 | 0.91 |
| *P*-value | < 0.001 | < 0.001 | < 0.001 | < 0.001 | < 0.001 |

| **Group** | **Urea** | **Calcium ion** | **Sodium** | **Chloride** |
|---|---|---|---|---|
| | **MTAC** | **MTAC** | **MTAC** | **MTAC** |
| Cumulative fill volume | $\mu L\ min^{-1}$ | $\mu L\ min^{-1}$ | $\mu L\ min^{-1}$ | $\mu L\ min^{-1}$ |
| **First dwell phase** | | | | |
| 8 mL | 200 (175 – 222) | 175 (113 – 233) | 140 (134 – 156) | 118 (96 – 133) |
| 12 mL | 208 (186 – 225) | 222 (206 – 248) | 170 (148 – 188) | 123 (92 – 160) |
| **Second dwell phase** | | | | |
| 8+8 mL | 331 (318 – 379) | 358 (292 – 473) | 248 (184 – 299) | 192 (168 – 210) |
| 12+12 mL | 430 (392 – 467) | 423 (355 – 463) | 297 (263 – 348) | 233 (202 – 309) |
| **Third dwell phase** | | | | |
| 8+8+8 mL | 405 (375 – 479) | 498 (403 – 736) | 248 (184 – 299) | 170 (139 – 201) |

| | | | | |
|---|---|---|---|---|
| 12+12+12 mL | 543 (515 – 591) | 657 (532 – 716) | 297 (263 – 348) | 266 (222 – 294) |
| **Spearman rank test** (IPV *vs* column) | | | | |
| Correlation coefficient (raw), *ρ* | 0.90 | 0.87 | 0.66 | 0.69 |
| Correlation coefficient (regularized), *ρ* | 0.91 | 0.91 | 0.86 | 0.88 |
| *P*-value | < 0.001 | < 0.001 | < 0.001 | < 0.001 |

**Supplemental Table 3.** Monte Carlo cross-validation results

| | Small solute diffusion capacity (MTAC) † | | | Osmotic conductance to glucose (OCG) | | |
|---|---|---|---|---|---|---|
| **Experimental data** | **RMSE** | **Break-point, mL** | **MTAC ‡** µL/min | **RMSE** | **Break-point, mL** | **OCG ‡** nL/min/mmHg |
| Square-Cube law | 52.1 (42.0 – 57.5) | - | 120 (116 – 124) | 9.1 (7.8 – 11.1) | - | 48 (47 – 48) |
| Break-point model | 50.8 (41.8 – 56.6) | 25.8 (25.2 – 26.4) | 149 (143 – 156) | 9.0 (7.8 – 11.0) | 27.8 (25.7 – 30.7) | 61 (59 – 65) |
| T-test, *p*-value | < 0.001 | | | < 0.001 | | |
| **Clinical data** | **RMSE** | **Break-point, mL** | **MTAC ‡** mL/min | **RMSE** | **Break-point, mL** | **OCG ‡** µL/min/mmHg |
| Square-Cube law | 0.22 (0.20 – 0.25) | - | 9.1 (8.9 – 9.3) | 1.007 (0.84 – 1.17) | - | 2.8 (2.5 – 3.2) |
| Break-point model | 0.21 (0.19 – 0.24) | 2,286 | 9.5 (9.2 – 9.8) | 1.005 (0.82 – 1.19) | 2,286 | 3.3 (2.9 – 3.7) |
| T- test, *p*-value | < 0.001 | | | < 0.001 | | |

RMSE, Root-mean-square error between model predictions and the validation datasets

† $^{51}$Cr-EDTA diffusion capacity (for experimental data), or creatinine diffusion capacity (clinical data)

‡ Value at the break-point or at 20/2000 mL (experimental data/clinical data).

**Supplemental Table 4.** Clinical scenarios simulated using the three-pore model

| Regimen | Dwell time | Initial intra-peritoneal volume | D/P creatinine | Trans-peritoneal UF volume † | Net UF ‡ |
|---|---|---|---|---|---|
| Conventional PET using 2.3% glucose | **240** | **2,300 mL** | **0.73** | **296 mL** | **224 mL** |
| Shorter PET using 2.3% glucose | **190** | **1,150 mL** | **0.74** | **146 mL** | **89 mL** |
| Shorter PET using 2.3% glucose | **151** | **575 mL** | **0.74** | **73 mL** | **27 mL** |

Dwell time for the shorter treatments were calculated by multiplying 240 min by $(1{,}150/2{,}300)^{1/3}$ and $(575/2{,}300)^{1/3}$, respectively.

† Net water transport across the peritoneal membrane (does not include reabsorption rate/lymphatic flow)

‡ Net water transport including a fixed reabsorption rate/lymphatic flow of 0.3 mL/min

**Calibration of analytical methods using the iSTAT-1**

*Sodium and chloride*

$Na^+$ and $Cl^-$ were measured with the CHEM8 cassette (Abbott, Abbott Park, IL) utilizing ion-selective electrode potentiometry as described the manufacturer. Bland-Altman analysis was performed using twenty-seven reference solutions, showing a variation coefficient (VC) of 0.8% for sodium and a VC 1.7% for chloride. There were small matrix effects for sodium when spiking with glucose as well as bicarbonate (both increased apparent sodium concentration). Correction was performed using:

$$\mathrm{f}^{-1} = \mathrm{a} + \mathrm{b} \cdot \mathrm{glu} + \mathrm{c} \cdot \mathrm{bic}$$

where **actual Na** = f × **measured Na** [a = 0.9815, b = $7.7 \cdot 10^{-5}$, c = $9.2 \cdot 10^{-4}$]. One can see that f equals ~1 for a glucose concentration (normal) of 5 mmol/L and a bicarbonate concentration (normal) of 25 mmol/L, illustrating that the iSTAT-1 device is calibrated for blood plasma measurements. There were matrix effects for chloride, also for glucose and bicarbonate (both led to the apparent chloride concentration being lower than actual, again **actual Cl** = f × **apparent Cl**:

$$\mathrm{f}^{-1} = \mathrm{a} + \mathrm{b} \cdot \mathrm{glu} + \mathrm{c} \cdot \mathrm{bic}$$

Here, a = 0.93, b = $-1.1 \cdot 10^{-4}$, c = $-1.5 \cdot 10^{-3}$.

*Bicarbonate*

Bicarbonate was estimated on the basis of the Henderson-Hasselbach equation from pH, $pCO_2$, and ionic strength ($Na^+$) measurements (according to the manufacturer, CHEM8 cassette), and calibrated to the International Federation of Clinical Chemistry (IFCC) $TCO_2$ reference method [1]. Bland-Altman analysis performed on the basis of twenty samples of standard fluids (reference) with known amounts of bicarbonate, lactate, and glucose concentrations revealed an imprecision (VC) of 7.5%. No matrix effects were detected for the included spike agents.

*Calcium ion*

Calcium ion was measured using the CHEM8 cassette. Twenty-seven samples of standard fluids having a known calcium ion concentration of 1.25 mmol/L with known amounts of bicarbonate, lactate and glucose concentrations were analyzed. Bland-Altman analysis showed an imprecision (VC) of 4.1%. A significant matrix effect was identified only for glucose, **actual Ca** = f × **apparent Ca**:

$$\mathrm{f}^{-1} = \mathrm{a} + \mathrm{b} \cdot \mathrm{glu}$$

Here, a= 0.74 and b=$2.3 \cdot 10^{-4}$.

### Segmented linear regression

The square-cube model and break-point models may be linearized:

$$\log y = \log\left(p_0\left(\frac{v}{v_t}\right)^a\right) = a\log\frac{v}{v_t} + \log p_0$$

Here, $y$ are the data points (i.e., OCG or MTAC values), $v$ is the intra-peritoneal volume, and $p_0$ is the value of the variable at the intra-peritoneal volume $v_t$ set to 2,286 mL for the square cube model. For the square-cube model, a=2/3, and for the break-point model, a=2/3 for the first segment (up to the break-point) and then a=1/3. The calculation of $p_0$ for the square-cube model is straightforward

$$\mathrm{p}_0 = e^{\frac{1}{N}\sum_{i=1}^{N}\log y_i - \frac{2}{3}\log\frac{v_i}{v_t}}$$

The root-mean-square error for the square-cube model is then calculated as

$$RMSE_0 = \frac{1}{N}\sum_{i=1}^{N}\left(\log y_i - \frac{2}{3}\log\frac{v_i}{v_t} - \log p_0\right)^2$$

Since the data points are distributed over 6 different intra-peritoneal volumes (688 mL, 1200 mL, 1694 mL, 2286 mL, 2817 mL and 3329 mL), there are four possible break-point models (one for each interior volume). The calculations of the 4 parameters $p_0$ is similar, but performed only over the segments A={688 mL, 1200 mL}, B={688 mL, 1200 mL, 1694 mL}, C={688 mL, 1200 mL, 1694 mL, 2286 mL}, D={688 mL, 1200 mL, 1694 mL, 2286 mL, 2817 mL}, as follows

$$\mathrm{p}_{0,\mathrm{A}} = e^{\frac{1}{2}\sum_{i\in A}\log y_i - a\log\frac{v_i}{v_t}}$$

$$\mathrm{p}_{0,\mathrm{B}} = e^{\frac{1}{3}\sum_{i\in B}\log y_i - a\log\frac{v_i}{v_t}}$$

$$\mathrm{p}_{0,\mathrm{C}} = e^{\frac{1}{4}\sum_{i\in C}\log y_i - a\log\frac{v_i}{v_t}}$$

$$\mathrm{p}_{0,\mathrm{D}} = e^{\frac{1}{5}\sum_{i\in D}\log y_i - a\log\frac{v_i}{v_t}}$$

The root-mean-square errors are then calculated as follows

$$RMSE_A = \frac{1}{N}\left(\sum_{i\in A}\left(\log y_i - \frac{2}{3}\log\frac{v_i}{v_t} - \log p_0\right)^2 + \sum_{i\notin A}\left(\log y_i - \frac{1}{3}\log\frac{v_i}{v_t} - \log p_0\right)^2\right)$$

$$RMSE_B = \frac{1}{N}\left(\sum_{i\in B}\left(\log y_i - \frac{2}{3}\log\frac{v_i}{v_t} - \log p_0\right)^2 + \sum_{i\notin B}\left(\log y_i - \frac{1}{3}\log\frac{v_i}{v_t} - \log p_0\right)^2\right)$$

$$RMSE_C = \frac{1}{N}\left(\sum_{i\in C}\left(\log y_i - \frac{2}{3}\log\frac{v_i}{v_t} - \log p_0\right)^2 + \sum_{i\notin C}\left(\log y_i - \frac{1}{3}\log\frac{v_i}{v_t} - \log p_0\right)^2\right)$$

$$RMSE_D = \frac{1}{N}\left(\sum_{i\in D}\left(\log y_i - \frac{2}{3}\log\frac{v_i}{v_t} - \log p_0\right)^2 + \sum_{i\notin D}\left(\log y_i - \frac{1}{3}\log\frac{v_i}{v_t} - \log p_0\right)^2\right)$$

Lastly, root-mean-squared errors had an apparent right-tailed distribution much like the log-normal distribution of which the first moment is the geometric mean. T-tests were therefore carried out on log-transformed RMSE.

### Mathematical derivation of dwell-time and UF factor

**Theorem 1:** Assume that the intra-peritoneal volume is changed from V to $V_{new}$, and that the dialysate concentration D of a solute species follows an exponential model $D(t)=D_0\exp(Vt/MTAC)$. Then, the time it takes to reach a dialysate concentration ratio $(D/D_0)$ using the new fill volume $V_{new}$ is reduced/increased by the factor $(V_{new}/V)^{1/3}$.

*Proof:* The time *T* it takes to reach a dialysate concentration D from an initial concentration $D_0$ is

$$T = \frac{V}{MTAC}\log\frac{D}{D_0}$$

Changing the volume from V to $V_{new}$, we get

$$T_{new} = \frac{V\left(\frac{V_{new}}{V}\right)}{MTAC\left(\frac{V_{new}}{V}\right)^{2/3}}\log\frac{D}{D_0} = \left(\frac{V_{new}}{V}\right)^{1/3}\frac{V}{MTAC}\log\frac{D}{D_0}$$

Thus, the time to reach $D/D_0$ is reduced/increased by the factor $(V_{new}/V)^{1/3}$, meaning that when the fill volume is decreased the patient will appear "faster" and *vice versa*.

**Theorem 2:** In addition to the assumptions in Theorem 1, assume that the osmotic gradient is maintained by adjusting the dwell time by the factor $(V_{new}/V)^{1/3}$. Then, the UF-rate is reduced/increased by the factor $(V_{new}/V)$.

Using Ohm's law [2], the UF rate is given by

$$UFR = OCG \cdot 19.3T(D_0 - D)/\log(D_0/D)$$

Again, changing the volume from V to $V_{new}$ and also changing the time to achieve the same osmotic gradient $(D/D_0)$, we get

$$UFR = OCG\left(\frac{V_{new}}{V}\right)^{2/3} \cdot 19.3T\left(\frac{V_{new}}{V}\right)^{1/3}(D_0 - D)/\log(D_0/D) \leftrightarrow$$

$$UFR = \left(\frac{V_{new}}{V}\right)OCG \cdot 19.3T(D_0 - D)/\log(D_0/D)$$

Thus, changing the volume from V to $V_{new}$, the UF-rate is decreased by the factor ($V_{new}/V$) if the dwell time is also adjusted by the factor $(V_{new}/V)^{1/3}$ to achieve a similar osmotic gradient.

6. Inc. APoC. *i-STAT 1 System Manual* Emergo Europe: The Hague, Netherlands, 2015.
